# A Multi-Omics Study Reveals Pathway-Level Insights and Predictive Biomarkers in pediatric TB

**DOI:** 10.64898/2026.03.13.26348257

**Authors:** Zaynab Mousavian, Mark R. Segal, Roger I. Calderon, Juaneta Luiz, Esin Nkereuwem, Peter Wambi, Mandar Paradkar, Molly F. Franke, Gunilla Källenius, Beate Kampmann, Aarti Kinikar, George B. Sigal, Christopher Sundling, Danielle L. Swaney, Eric Wobudeya, Heather J. Zar, Jeffrey M. Collins, Adithya Cattamanchi, Joel D. Ernst, Devan Jaganath

## Abstract

**Background:** Tuberculosis (TB) remains a global health threat, affecting over a million children under the age of 15 annually. Many children with TB do not receive treatment due to challenges in diagnosis.

**Methods:** We performed a multi-omics analysis for pediatric TB by integrating plasma proteomics and metabolomics data from children with presumptive TB across four high-burden countries. Pathway enrichment analysis was conducted using multiGSEA to identify relevant immune and metabolic pathways. We also applied mixOmics and multiview approaches for diagnostic biomarker discovery and compared the performance of multi-omics signatures with those derived from single-omics datasets.

**Results:** Enrichment analysis revealed several immune and metabolic pathways, including PTEN and RUNX2 regulation pathways, as well as arginine and proline metabolism, that were uniquely identified through data integration. While the multi-omics model showed marginal improvement over single-omics models, proteomics alone generally outperformed metabolomics and demonstrated greater potential for accurately classifying Confirmed TB versus Unlikely TB in children.

**Conclusion:** These findings demonstrate the advantage of combining complementary molecular layers to gain a deeper understanding of disease mechanisms and highlight the potential of proteomics for improving pediatric TB diagnosis.

## 1. Introduction

Tuberculosis (TB) remains a leading cause of death from infectious diseases, with over 1 million children under 15 years falling ill each year [1]. More than half of these cases are not reported to public health programs in part due to the lack of sensitive diagnostic tests and limited diagnostic capacity in the healthcare settings where these individuals seek care [2]. Diagnosing TB in children is particularly challenging due to non-specific symptoms, lower bacterial loads, and difficulties obtaining sputum samples; factors that render many conventional diagnostic tools ineffective [3]. Furthermore, TB in children has been studied less extensively compared to TB in adults [4], resulting in a limited understanding of the disease’s underlying mechanisms in this age group.

Advancements in omics technologies over the last decades have enabled systems-level insights to better understand the mechanisms of TB and potentially identify novel molecular signatures of the disease. To date, there has been a growing number of studies on TB biomarker discovery in children [5], but most have focused on transcriptomics, proteomics, or metabolomics alone or have profiled only a small number of hypothesis-driven transcripts, proteins, or metabolites, rather than using discovery-based approaches. Moreover, no multi-omics studies have focused on childhood TB, potentially overlooking biomarkers or biological pathways that are only discernible through integrated data analysis.

Building on our previous studies that analyzed plasma proteomics [6] and metabolomics data [7] separately to identify TB biomarkers in children, we performed an integrated multi-omics analysis in the subset of participants where both proteomic and metabolomic data were available. Although the same underlying cohort framework and previously generated datasets were used, as described in the original studies [6, 7], this study integrates the plasma proteomics and metabolomics data from these participants. By expanding our analysis to include multiple data types across a larger number of biologic pathways, we sought to gain new insights into the host to TB in children. As a secondary objective, we aimed to leverage integrative multi-omics methods for biomarker discovery to evaluate whether a combined protein and metabolite signature could identify TB with higher accuracy compared to each data type alone.

## 2. Materials and Methods

### 2.1. Pediatric TB cohort

The plasma samples used in this study were originally collected as part of prospective pediatric TB studies [6, 7] conducted at four international sites: The Gambia, Peru, South Africa, and Uganda (Supplementary Figure 1). Children under 15 years of age were enrolled if they were being evaluated for pulmonary TB disease. Eligibility criteria included either microbiologically confirmed TB (via Xpert MTB/RIF, Xpert Ultra, or culture) or clinical suspicion of TB based on unexplained cough of any duration and at least one additional symptom or sign suggestive of intrathoracic TB (including weight loss and poor weight gain/failure to thrive; persistent fever for longer than one week, unexplained lethargy or reduced playfulness for more than one week, abnormal chest X-ray, positive tuberculin skin test (TST), or known TB contact). Detailed information on study design, eligibility, and procedures has been previously published [6, 7].

All participants provided written informed consent for future sample use. Ethical approval was obtained from the institutional review boards at all participating sites including the Mulago Hospital Ethics Research Committee, Gambian Government and MRC Joint Ethics Committee, London School of Hygiene and Tropical Medicine, Institutional Ethics Committee for Research of the National Institute of Health – Peru, Human Research Ethics Committee of the Faculty of Health Sciences, University of Cape Town, and the University of California, San Francisco (UCSF) IRB. For this analysis, we focused on children with clearly defined TB status: those with microbiologically confirmed TB (Confirmed TB) and those without clinical or microbiologic evidence of TB (Unlikely TB). To avoid misclassification (i.e., the inclusion of false positive TB cases), we excluded children with Unconfirmed TB, defined as those with clinical signs suggestive of TB but without microbiologic confirmation. Study groups were defined using standard terminology from the National Institutes of Health (NIH) classification for childhood TB [6, 7], and all participants met the corresponding criteria.

### 2.2. Protein and metabolite abundance data analysis

Proteomic and metabolomic data analyzed in this study were generated previously using mass spectrometry, as reported in [6, 7]. We used the *lmFit* and *eBayes* functions from the limma R package [8] to determine the fold change, and attendant p-values (adjusted for multiple testing using the Benjamin-Hochberg False Discovery Rates [9] where applicable), of protein and metabolite abundance levels between Confirmed TB and Unlikely TB groups. Both site and age group were included as covariates in the analysis of differential proteins and metabolites. To remove site-specific effects, the *removebatcheffec*t function from the limma package was applied to both datasets. To demonstrate the effect of batch correction, we performed Principal Component Analysis (PCA) using the *prcomp* function in R.

### 2.3. Multi-omics pathway enrichment analysis

For pathway enrichment analysis, we used the multiGSEA R package [10] to identify integrated metabolomic and proteomic pathway activity in TB. We used the fold change values and p-values obtained from the differential expression analysis (DEA) for each protein and metabolite as input for the multiGSEA method. In this approach, gene set enrichment analysis (GSEA) is performed separately on the proteomics and metabolomics dataset. To integrate findings from each dataset, we aggregated p-values using Stouffer’s Z-method as implemented in the multiGSEA package. We utilized both the KEGG [11] and REACTOME [12] databases as resources for signaling pathways.

### 2.4. Multi-omics biomarker discovery analysis

To integrate proteomics and metabolomics data for biomarker discovery, we used two different R packages: mixOmics [13] and multiview [14]. In mixOmics, we applied the Data Integration Analysis for Biomarker discovery using Latent cOmponents (DIABLO) method [15] for data integration. This method identifies latent components that best capture the variation in the data across different omics layers. The sparse partial least squares (sPLS) analysis used in DIABLO acts as a regularization technique to prevent overfitting and assist in feature selection. In this method, the loading factor represents the weight or contribution of each variable (e.g., a protein or metabolite) to a given latent component, helping to identify the features most influential in the integrated model. In the multiview package, we employed the Least Absolute Shrinkage and Selection Operator (LASSO) method [16], considering both proteomics and metabolomics datasets simultaneously. We tested different values of the weighting parameter (rho), ranging from 0 to 1. A value of 0 corresponds to combining the proteomics and metabolomics features before training the model (early fusion), while a value of 1 corresponds to combining the predictions from separate models trained on each data type (late fusion). To identify the features that yielded good predictive accuracy, we utilized two conventional means for model size determination according to differing degrees of penalization as governed by the regularization parameter (lambda: lambda.min and lambda.1se). Accuracy for the multi-view model was assessed in two ways: misclassification rate and AUC.

To assess the performance of each signature identified by different methods, we used a support vector machine (SVM) [17] with a radial basis function (RBF) kernel, implemented via the svmRadial method in the caret R package [18]. This model is well-suited to capture heterogeneous and potentially non-linear relationships commonly observed in multi-omics datasets. The area under the receiver operating characteristic curve (AUC-ROC) was calculated for each model, along with 95% confidence intervals (CIs), to evaluate overall classification performance. The ROC curves illustrate the sensitivity and specificity of each signature in distinguishing Confirmed TB from Unlikely TB across various thresholds. To compare each multi-omics model with the proteomics and metabolomics models, we used features from both omics layers to train the classification model. In our previous proteomics study [6], we found that individual markers had worse classification accuracy for identifying TB in children than signatures that included multiple proteins. Therefore, in this study, we focused on identifying improved marker combinations through multi-omics integration and did not evaluate single-marker performance.

## 3. Results

### 3.1. Clinical characteristics of the study participants

We included 237 plasma samples from children enrolled at four clinical sites: The Gambia (n=77), Peru (n=49), South Africa (n=37) and Uganda (n=74) (Table 1). This analysis consisted of 80 samples with Confirmed TB and 157 samples classified as Unlikely TB (Figure 1). The median age was 4 years (IQR 2-7) for those with Confirmed TB, and 5 years (IQR 2-8) for those with Unlikely TB. There were 114 female and 123 male children, with approximately equal numbers of females and males in each group based on TB status. Out of 237 children, 18 (8%) were living with HIV, equally distributed between the Confirmed TB and Unlikely TB groups, with 13 from South Africa and five from Uganda. The weight-for-age z-score (WAZ) was below -2 for about half of the children with Confirmed TB in The Gambia and South Africa, indicating a high prevalence of malnutrition in the Confirmed TB group within these two cohorts.

**Table 1.** Overview of demographic and clinical characteristics of the study cohort.

| Characteristics | Confirmed TB<br>(N=80) | Unlikely TB<br>(N=157) | Total<br>(N=237) |
| --- | --- | --- | --- |
| The Gambia | 29 (38%) | 48 (62%) | 77 |
| Peru | 17 (35%) | 32 (65%) | 49 |
| South Africa | 18 (49%) | 19 (51%) | 37 |
| Uganda | 16 (22%) | 58 (78%) | 74 |
| Age median (IQR) | 4 (2-7) | 5 (2-8) | 4 (2-8) |
| Female N (%) | 32 (40%) | 82 (52%) | 114 (48%) |
| HIV positive, N (%) | 8 (10%) | 10 (6%) | 18 (7%) |
| WAZ<-2, N (%) | 18 (22%) | 14 (9%) | 32 (13%) |

**Figure 1.**
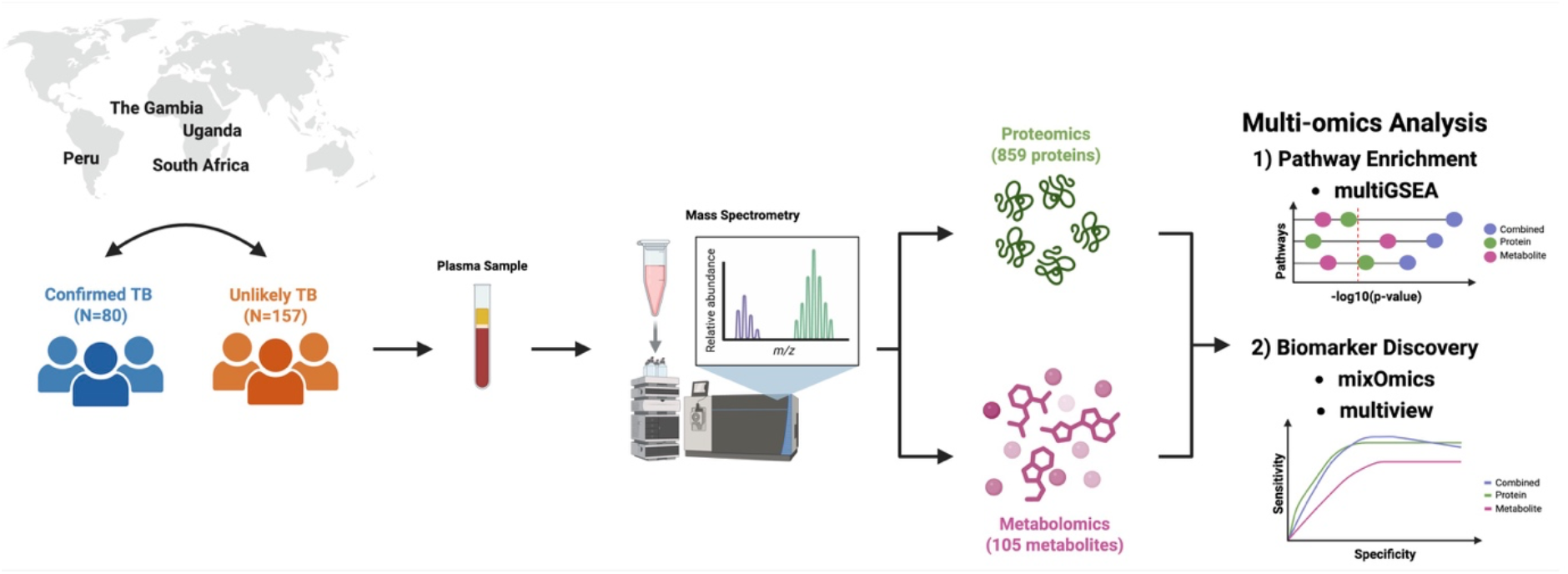
A schematic representation of the study plan.

### 3.2. Pathways significantly enriched in Confirmed TB vs Unlikely TB

Our dataset consisted of abundance levels for 859 proteins and 105 metabolites across all participants, which have been previously published [6, 7]. Performing principal component analysis (PCA) on the protein and metabolite data separately, we observed a clear separation between samples from Peru and the other sites for the protein dataset (Supplementary Figure 2a), while samples from The Gambia were clearly separated from the other samples in the metabolite dataset (Supplementary Figure 2c). We removed these site-specific effects and ensured that the data distribution between groups remained unchanged following this preprocessing (Supplementary Figures 2b and 2d).

Our first aim was to understand which pathways are enriched in Confirmed TB versus Unlikely TB in pediatric patients. After conducting differential expression analysis (DEA) on each dataset (Supplementary Figures 3a and 3b), out of 2,799 pathways collected from KEGG and REACTOME databases, 115 and 74 were enriched in Confirmed TB versus Unlikely TB (p-value < 0.05) using the DEA results from the proteomics and the metabolomics datasets, respectively (Figure 2). Combining p-values from both datasets, we identified 42 signaling pathways significantly enriched (combined p-value<0.05) in Confirmed TB compared to Unlikely TB (Figure 2); however, none of the pathways were enriched based on the BH FDR– corrected p-value. Among them, only a small subset (seven pathways) showed significant enrichment in both the proteomics and metabolomics dataset. The seven shared pathways included those related to the cell cycle, viral infection, and SARS-CoV infection. Since all participants included in this study were recruited prior to the COVID-19 pandemic, the enrichment of pathways annotated as viral infection or SARS-CoV–related reflects shared host immune and inflammatory signaling mechanisms rather than true COVID-19 exposure. These pathways capture conserved processes such as immune system, innate immune activation, and cellular response to stress that are common to both TB and viral infections.

**Figure 2.**
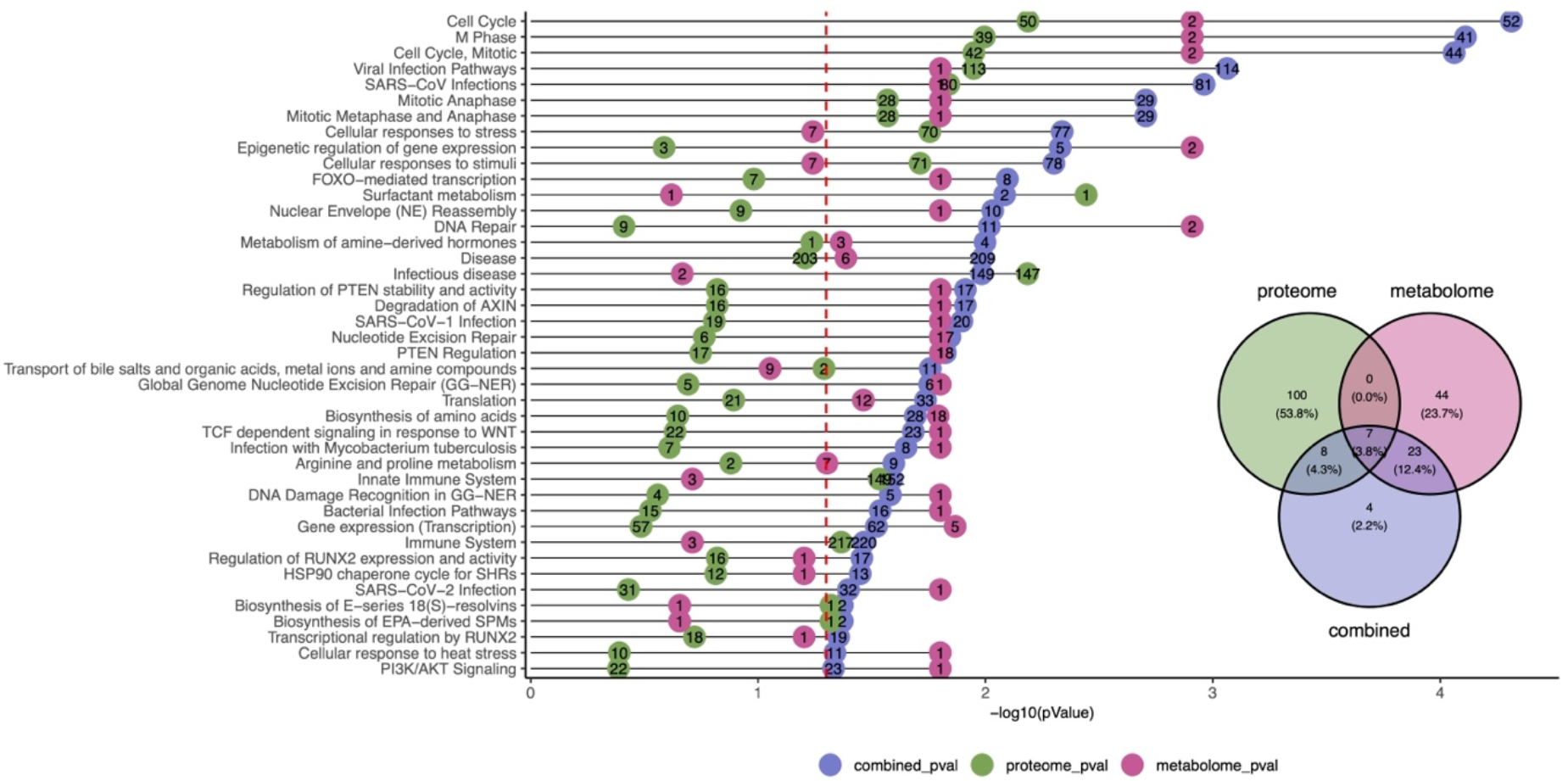
Pathway enrichment analysis using the multiGSEA approach. Each row represents a signaling pathway with a combined p-value < 0.05. The dashed red line shows the significance threshold of 0.05. Purple, green, and pink bars indicate p-values from the combined analysis, proteomics dataset, and metabolomics dataset, respectively. Numbers on the bars represent the number of overlapping proteins or metabolites between our dataset and each pathway. The Venn diagram indicates the number and percentage of pathways identified by each omics layer and the integrated analysis.

Moreover, four pathways, including regulation of RUNX2 expression and activity, transcriptional regulation by RUNX2, HSP90 chaperon cycle for SHRs, and arginine and proline metabolism were enriched only when results from both datasets were integrated. Among the pathways enriched in the integrated analysis, most (27) were not found in the list of pathways enriched in the proteomics dataset, while 12 were not found in the list of enriched pathways in the metabolite dataset. Among those 27 pathways which were not enriched in the proteomics dataset alone, we observed the bacterial infection pathways, infection with Mycobacterium tuberculosis, and PTEN regulation. Our analysis revealed that the metabolite nicotinamide plays a central role in the enrichment of multiple pathways, as it was upregulated in Confirmed TB versus Unlikely TB in the metabolomics dataset.

### 3.3. Multi-omics TB signature biomarker discovery

As a second aim, we examined both mixOmics and multiview methods to integrate protein data with metabolite data and determined whether a multi-omics signature improved classification accuracy for TB in children versus either method alone. Before proceeding with further analyses, 40% of the entire dataset was reserved for independent validation of the selected signatures to ensure the robustness and generalizability of the classification models, and the remaining 60% was used for supervised discovery and training. Both training and test datasets had the same proportional representation from each clinical group and study site.

Using the DIABLO approach, we identified two components from the proteomics dataset and two components from the metabolomics dataset for classification of Confirmed TB versus Unlikely TB. Proteins and metabolites with the highest absolute loading factors indicated the features that contributed most to the latent components in the DIABLO model. In each component from the proteomics dataset, 10 different proteins contributed with varying loading factors (Supplementary Figures 4a and 4b). As indicated in Supplementary Figure 5, five proteins were highly correlated (i.e., r>0.6) with each component. These included APOM, CRP, WARS1, IGFALS, and GPLD1 for component 1 and APOA2, ITIH1, CD5L, CBLN4, and PLG for component 2. Among them, four proteins (APOM, CRP, APOA2, and ITIH1) had an absolute loading factor above 0.4 (Supplementary Figures 4a and 4b). For the metabolomics dataset, each component included five contributing metabolites with varying loadings (Supplementary Figures 4c and 4d). Four metabolites (retinol, tryptophan, citrulline, and hydroxyphenylpropionate) and three others (methionine sulfoxide, arginine, and histamine) were highly correlated with components 1 and 2, respectively (Supplementary Figure 5). Among these, four metabolites (hydroxyphenylpropionate, tryptophan, methionine sulfoxide, and histamine) had an absolute loading factor above 0.4 (Supplementary Figures 4c and 4d). For multi-omics classification, we constructed three types of SVM models, protein-only, metabolite-only, and a combined protein–metabolite model, using three different feature sets: (1) all contributing proteins and metabolites to the identified components in the DIABLO model, (2) a subset of variables highly correlated with the components identified in the prior analysis, and (3) a subset consisting only of those with high loading factors (i.e., absolute loading factor > 0.4). While performance varied slightly across the three feature sets, the third feature set yielded the highest AUC for the combined model (AUC = 0.73, 95% CI: 0.62–0.83), with AUCs of 0.67 for the protein model and 0.54 for the metabolite model (Figure 3a). This feature set included the proteins APOM, CRP, APOA2, and ITIH1, and the metabolites hydroxyphenylpropionate, tryptophan, methionine sulfoxide, and histamine. The ROC curves for the other two feature sets are provided in Supplementary Figure 6.

**Figure 3.**
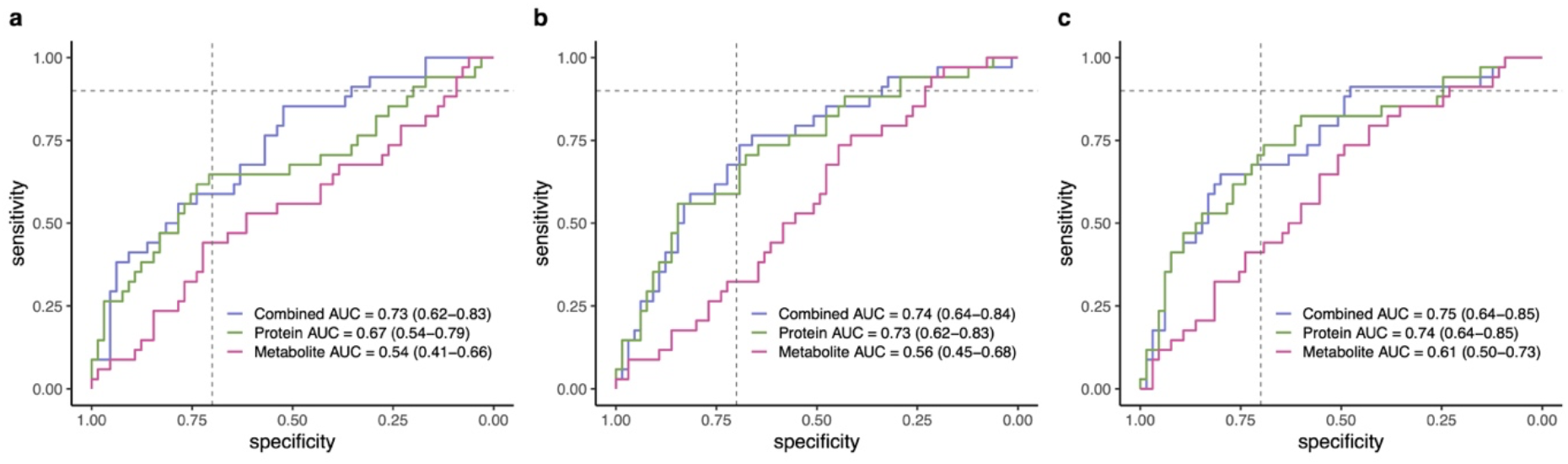
Classification performance of models using different feature sets for classifying Confirmed TB versus Unlikely TB (proteins in green, metabolites in pink, and combined in purple): (a) ROC curves for models using high-loading proteins and metabolites from mixOmics (4 proteins and 4 metabolites) (b) ROC curves for models using a minimal set of features selected from multiview (2 proteins and 3 metabolites). (c) ROC curves for the models using the combined features (5 proteins and 6 metabolites) from both mixOmics and multiview analysis. AUC values with 95% confidence intervals are included for each model. The two dashed lines in two plots represent the target product profile (TPP) criteria for a TB triage test, defined by a sensitivity above 90% and a minimum specificity of 70%.

As a second multi-omics biomarker discovery approach, the multiview model was used and trained using two strategies: one based on minimizing misclassification and the other optimized directly for the AUC metric. We explored a range of rho values and evaluated model performance based on AUC, sensitivity at 70% specificity, and the number of selected features. This evaluation allowed us to identify models that achieved the best classification performance while maintaining feature parsimony (Supplementary Figure 7). Among models with fewer than 20 features, the protein model trained using misclassification with lambda.1se and rho = 0.1, including 19 proteins, achieved the highest AUC (AUC = 0.75). For metabolites, the model trained using AUC with lambda.min and rho = 0.1, including 18 metabolites, had the highest AUC (AUC = 0.61) (Supplementary Figure 7). Combining these two feature sets yielded an AUC of 0.76 (Supplementary Figure 8). Considering minimal feature sets (i.e., fewer than five features; Supplementary Figure 7), comparable performance was achieved (AUC = 0.74, 95% CI: 0.64–0.84) using a minimal subset comprising two proteins (APOM and SFTPB) and three metabolites (pyruvate, carnitine, and histamine) (Figure 3b). In comparison, models trained on proteomics or metabolomics data alone achieved AUCs of 0.73 and 0.56, respectively.

Using four proteins (APOM, CRP, APOA2, and ITIH1) from the DIABLO model in mixOmics (Supplementary Figures 4a and 4b) and two proteins (APOM and SFTPB) from the multiview model, we obtained a total of five unique proteins. Similarly, using four metabolites (hydroxyphenylpropionate, tryptophan, methionine sulfoxide, and histamine) from mixOmics (Supplementary Figures 4c and 4d) and three metabolites (pyruvate, carnitine, and histamine) from the multiview model, we had six unique metabolites. These were used for an additional ROC analysis (Figure 3c). The combined model showed a slight improvement, with an AUC of 0.75 (95% CI: 0.64-0.85). For individual omics datasets, the AUC for proteins was 0.74, while for metabolites, it was 0.61. These results consistently showed that proteins provided better predictive performance than metabolites. The integration of both protein and metabolite features resulted in only a slight improvement, indicating that combining the two datasets did not significantly enhance performance compared to using proteins alone.

## 4. Discussion

We conducted a multi-omics approach to pediatric TB by integrating proteomics and metabolomics data. Our primary goal was to evaluate whether combining these complementary datasets could enhance our biological understanding of TB. Pathway enrichment analysis demonstrated that integrating multi-omics data can reveal biological pathways not captured by individual omics datasets. Specifically, several immune response and metabolic pathways were uniquely identified through the integrated analysis, which highlights the importance of leveraging signals across molecular layers to uncover complex disease mechanisms. Our secondary goal was to assess if a multi-omics signature improved the accuracy to detect childhood TB compared to proteomics or metabolomics alone. While the accuracy was higher in the multi-omics signature compared to metabolomics alone, this was largely driven by the protein markers and there was only a modest increase in accuracy compared to the proteomic data alone. Our findings provide a greater understanding of the host pathways that are involved in TB pathogenesis for children, while guiding an optimal approach to pediatric TB biosignature development.

One of these pathways, regulation of RUNX2 expression and activity, has previously been linked to TB in a study demonstrating activation of the BMP/SMAD/RUNX2 pathway in Mtb-infected macrophages [19]. Additionally, another study identified RUNX2 as a novel regulator of airway goblet cell differentiation and the associated mucus overproduction, which are key processes in the pathogenesis of respiratory diseases such as asthma [20]. Moreover, arginine and proline metabolism was another enriched pathway in Confirmed TB with a significant combined p-value, showing a substantial overlap with our metabolite dataset, including seven shared metabolites. This pathway was also among the most significantly enriched in children with exacerbating asthma hospitalized for status asthmaticus [21], and has been identified as a potential source of biomarkers for exacerbation-prone asthma in children [22]. Arginine metabolism has been associated with asthma in children across multiple studies [23-25] and its dysregulation has been implicated in the pathogenesis of various pulmonary inflammatory diseases, including respiratory infections, asthma, COPD, and pulmonary fibrosis [26].

In addition, PTEN regulation emerged among these pathways and has been previously linked to susceptibility to mycobacterial infections [27]. PTEN is a lipid phosphatase best known for its tumor suppressor function and for modulating host cell resistance to intracellular pathogens by regulating the PI3K/Akt signaling axis [27, 28]. Studies have shown that PTEN deficiency increases cellular susceptibility to Mycobacterium species, suggesting that the PTEN pathway may influence host-pathogen interactions and contribute to immune evasion mechanisms in TB [27]. This suggests that multi-omics integration provides a more comprehensive picture of the host response to TB infection in children.

Our analysis also revealed the cell cycle pathway as the most enriched in Confirmed TB, consistent with evidence from pediatric respiratory infections such as Respiratory syncytial virus (RSV), where virus-induced cell cycle arrest in epithelial cells promotes viral replication via TGF-β signaling [29]. Although TB is bacterial, this convergence suggests that cell cycle dysregulation may represent a common mechanism linking pathogen-induced host responses to respiratory disease pathogenesis.

In this context, our metabolomics dataset showed that nicotinamide was significantly elevated in Confirmed TB compared with Unlikely TB and played a key role in driving the enrichment of several metabolic pathways. As the main NAD^+^ precursor, nicotinamide is central to cellular metabolism, energy production, and redox balance, and has been studied for its roles in DNA repair, stress responses, and anticancer mechanisms [30]. Nicotinamide (a form of vitamin B3) has a long history of investigation for its effects on *Mycobacterium tuberculosis* and host responses, with early research exploring its antimicrobial potential in humans [31]. Although its direct antimycobacterial activity in culture is generally modest, recent studies indicate that nicotinamide can modulate macrophage function to influence intracellular bacterial control [32]. Thus, the elevation of nicotinamide observed in Confirmed TB in our study aligns with these mechanisms and underscores its potential relevance as both a metabolic biomarker and a mediator of altered host metabolic responses during TB disease.

For biomarker discovery, proteomics consistently outperformed metabolomics across different analysis strategies. Classification models based on proteins alone achieved higher AUC values (AUC=0.75) compared to models based solely on metabolites (AUC=0.61). Although combining proteomics and metabolomics data sometimes led to marginal improvements in classification performance, the overall gain was limited. Even a small subset of proteins, such as APOM and SFTPB, showed very good predictive performance. This result suggests that adding metabolites to proteins provided limited improvement in diagnostic performance for pediatric TB. While our previous study [6] showed higher performance for a plasma protein biomarker, the reduced performance observed here may be due to the smaller dataset limited to samples with both proteomic and metabolomic data. Moreover, we used multi-omics approaches here to identify the optimal combination of markers from each dataset. Although multi-omics technologies have advanced rapidly, becoming more affordable and even enabling single-cell resolution, there remains significant potential to develop computational methods that can effectively integrate these data and identify the most predictive markers.

While several studies have explored multi-omics integration for TB diagnosis in adults [33], to our knowledge, this is the first study to apply such a multi-omics approach specifically to pediatric TB. For example, Krishnan et al. [34] identified hsa-miR-215-5p and gamma-glutamylthreonine as key classifiers of incident TB in people living with HIV, achieving high accuracy (AUC=0.965), though no metabolites were differentially abundant overall. Fernandes et al. [35] reported a proteomic and metabolomic signature that perfectly classified TB patients from healthy controls (AUC = 1), while Wang et al. [36] identified urinary biomarkers that diagnosed TB from healthy controls with approximately 86% sensitivity and 88% specificity. Pedersen et al. [37] found a plasma biosignature combining proteins and miRNAs that distinguished active TB from healthy controls with very good accuracy (AUC = 0.995), also showing markers that tracked treatment response. These prior adult-focused studies had healthy control groups which likely overestimated the accuracy of these host markers. In our study, the control group comprised children with non-TB respiratory illnesses, making the study population more representative of clinical settings where children are evaluated for TB disease.

One limitation of this analysis is the relatively low proportion of proteins and metabolites in each signaling pathway that were measured in our datasets. Nonetheless, the association of several identified pathways with the immune system and infectious diseases supports the overall validity and relevance of our findings. Future studies with broader molecular coverage or additional omics layers could help overcome this limitation. Moreover, none of the pathways identified in the pathway enrichment analysis remained statistically significant after correction for multiple hypothesis testing. Future studies with larger sample sizes and independent validation cohorts will be required to confirm these observations.

A high proportion of children with Confirmed TB in the study cohort were affected by undernutrition, a condition known to impact immune function and alter metabolic profiles [38]. While this reflects the real-world context of pediatric TB populations, where undernutrition is highly prevalent, and thus enhances the generalizability of our findings, it may also create high variability in metabolite levels and attenuate signals. However, when we compared median metabolite levels across nutritional status categories, we did not observe any metabolites showing significant differences. Moreover, due to the limited sample size, we were unable to perform subgroup analyses based on nutritional status, which could have provided further insight into how this influences biomarkers expression. Future studies with larger cohorts will be essential to evaluate the identified biomarkers across diverse clinical subgroups.

In summary, our study aimed to determine whether integrating multi-omics data could improve TB classification beyond single-omics measurements. We found that adding metabolites to protein data provided limited improvement, suggesting that proteomics alone captured most of the predictive signal. Nevertheless, multi-omics integration offered additional insights into disease mechanisms that would not have been captured by single-omics analysis alone. Future studies incorporating layers such as transcriptomics or immune profiling may provide a more comprehensive understanding and further enhance predictive performance.

## Author Contributions

Conceptualization: ZM, MRS, JMC, DJ, AC, JDE. Data curation: ZM, MRS, JMC, DJ. Formal analysis: ZM. Funding acquisition: AC, JDE, GK, CS, JMC, DJ. Investigation: RIC, JL, EN, PW, MP, MFF, BK, AK, EW, HJZ, ZM, JMC, DLS, DJ, AC, JDE. Methodology: ZM, MRS, JMC, DLS, DJ. Project administration: MFF, BK, AK, EW, HJZ, AC, JDE, JMC, DJ. Software: ZM. Resources: MFF, BK, AK, EW, HJZ, AC, JDE, JMC, DJ. Supervision: JMC, DJ. Validation: ZM, MRS, JMC, DJ. Visualization: ZM. Writing – original draft: ZM, JMC, DJ. Writing – review & editing: all authors.

## Acknowledgement

The authors thank the physicians, nurses, and staff at the participating clinical sites for their care of the children in this study. We are especially grateful to the children and their families for their willingness to participate and contribute valuable data that may benefit future patients with the same illness. The first author gratefully acknowledges the support of the Erik and Edith Fernström Foundation in Sweden and the VALIDATE Network for funding her six-month visiting program at UCSF, which made the completion of this study possible.

## Data Availability Statement

Data is provided within the manuscript or supplementary material. All code used in the analyses is deposited on https://github.com/zmousavian/COMBO-study. Any additional information required to reanalyze the data reported in this paper is available from the lead contacts upon request.

