## Supplementary Materials for "A Multi-Omics Study Reveals Pathway-Level Insights and Predictive Biomarkers in pediatric TB"

[1]Department of Global Health, Rollins School of Public Health, Emory University, Atlanta, GA, USA
[2]Department of Medicine, Division of Experimental Medicine, University of California San Francisco, San Francisco, CA, USA
[3]Division of Infectious Diseases, Department of Medicine Solna and Center for Molecular Medicine, Karolinska Institutet, Stockholm, Sweden
[4]Department of Epidemiology and Biostatistics, University of California San Francisco, San Francisco, CA, USA
[5]Advanced Research and Health, Lima, Peru
[6]Department of Pediatrics and Child Health, South African Medical Research Council Unit on Child and Adolescent Health, University of Cape Town, Cape Town, South Africa
[7]Department of Pediatrics, Dora Nginza Hospital, Gqeberha, South Africa
[8]Vaccines and Immunity Theme, MRC Unit The Gambia at the London School of Hygiene and Tropical Medicine, Fajara, The Gambia
[9]Uganda Tuberculosis Implementation Research Consortium, Walimu, Kololo, Kampala, Uganda
[10]Byramjee Jeejeebhoy Government Medical College-Johns Hopkins University Clinical Research Site
[11]Harvard Medical School
[12]Department of Infectious Diseases, Karolinska University Hospital, Stockholm, Sweden
[13]Charité Center for Global Health, Charité Universitätsmedizin Berlin, Berlin, Germany
[14]Byramjee Jeejeebhoy Government Medical College and Sassoon General Hospitals
[15]Meso Scale Diagnostics, LLC., Rockville, MD, USA
[16]J. David Gladstone Institutes, San Francisco, CA, USA
[17]Quantitative Biosciences Institute (QBI), University of California San Francisco, San Francisco, CA, USA
[18]Department of Cellular and Molecular Pharmacology, University of California San Francisco, San Francisco, CA, USA
[19]Division of Infectious Diseases, Department of Medicine, Emory University School of Medicine, Atlanta, GA, USA
[20]Institute for Global Health Sciences, Center for Tuberculosis, University of California San Francisco, San Francisco, CA, USA
[21]Division of Pulmonary Diseases and Critical Care Medicine, Department of Medicine, University of California Irvine, Irvine, CA, USA
[22]Department of Pediatrics, Division of Pediatric Infectious Diseases, University of California San Francisco, San Francisco, CA, USA

*** Correspondence:**
Zaynab Mousavian


Devan Jaganath


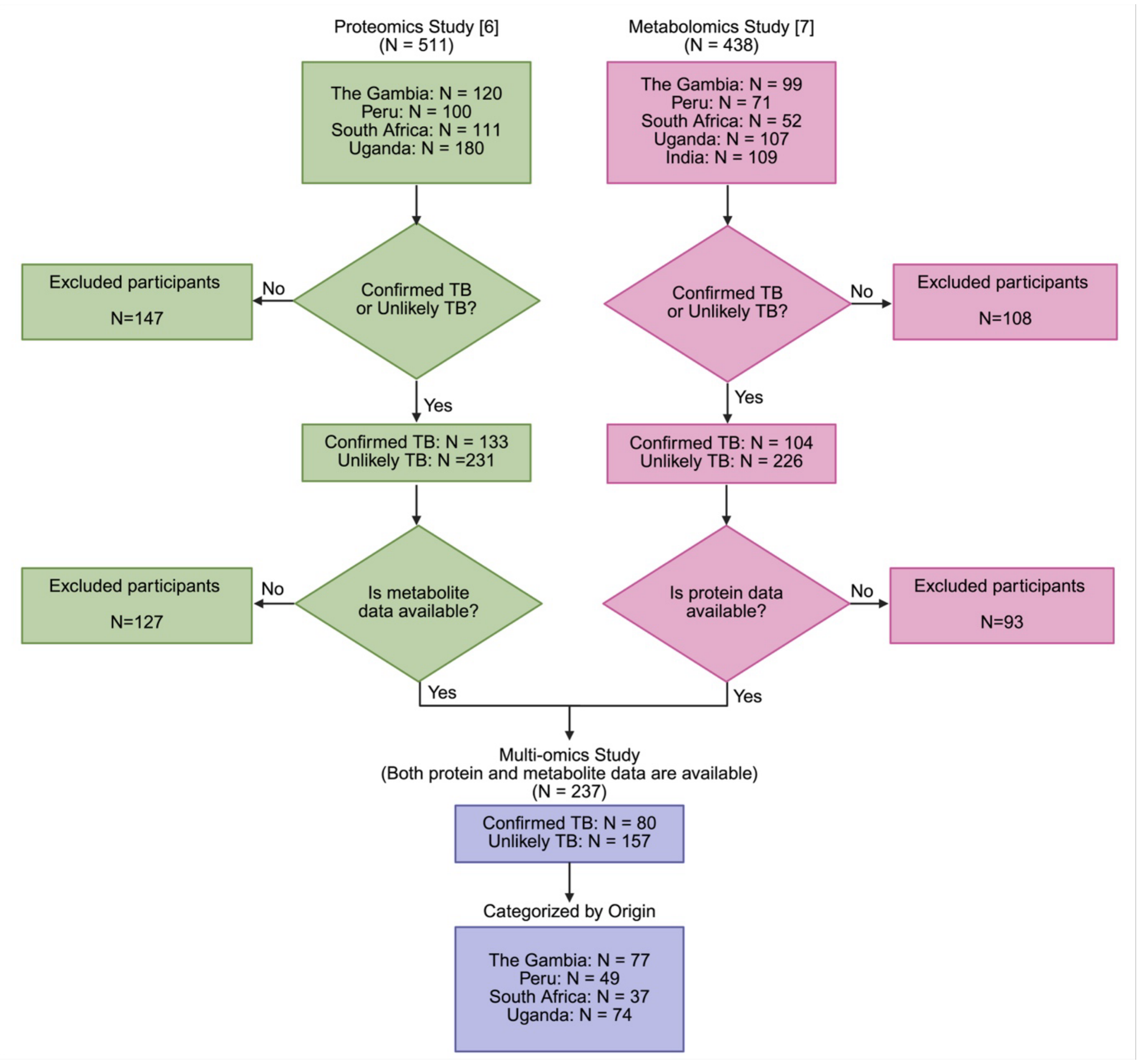


***Supplementary Figure 1.*** Participant selection flowchart for the multiomics study based on proteomics and metabolomics data.

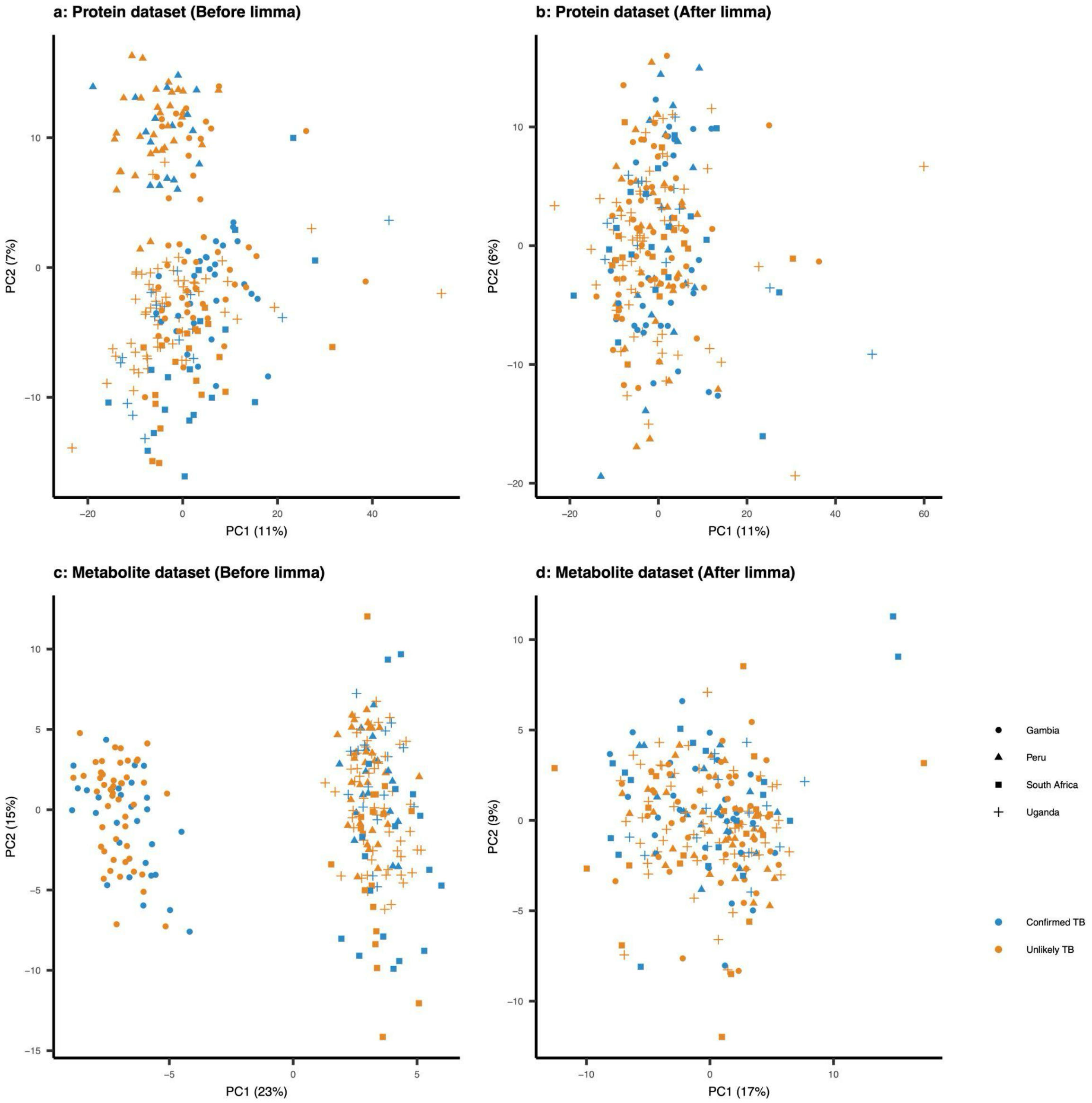


***Supplementary Figure 2.*** PCA plots of individuals in Confirmed TB (blue) and Unlikely TB (orange) groups based on proteomics and metabolomics data from four clinical sites: the Gambia (circle), Peru (triangle), South Africa (square), and Uganda (plus sign). Panels a and b represent the PCA plots before and after applying the removebatcheffect function from the limma R package to the proteomics data, while panels c and d show the corresponding plots for the metabolomics data. In each plot, the X-axis (PC1) and the Y-axis (PC2) represent principal components 1 and 2, respectively, with the percentage of variance in the dataset captured by each component (%).

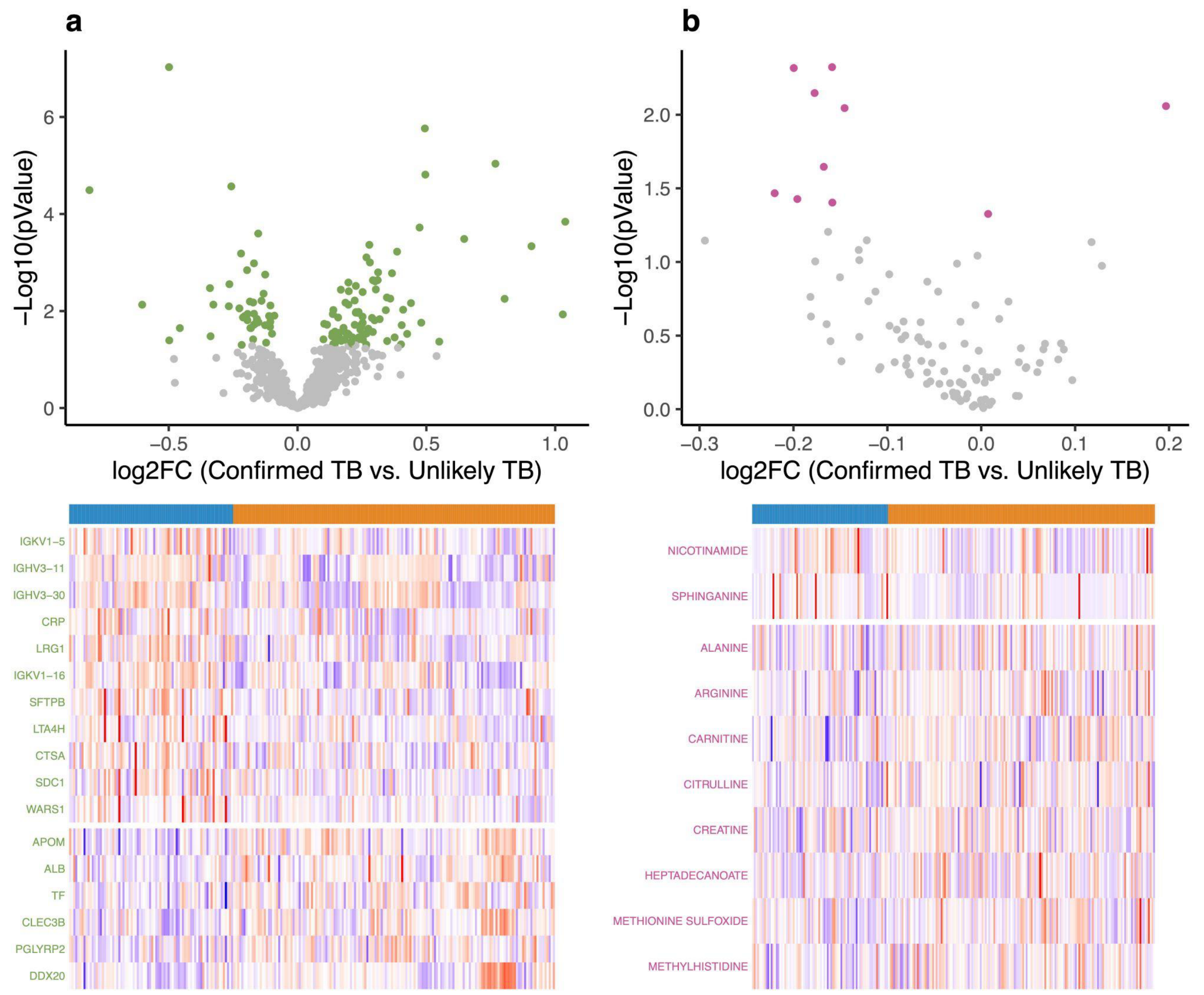


***Supplementary Figure 3.*** Differential analysis of proteomics and metabolomics data. (a) Volcano plot of proteomics data; proteins with p-value < 0.05 are shown in green, and Heatmap of significantly differentially abundant proteins between Confirmed TB (blue) and Unlikely TB (orange) groups (FDR-adjusted p-value < 0.05). (b) Volcano plot of metabolomics data; metabolites with p-value < 0.05 are shown in pink, and Heatmap of significantly differentially abundant metabolites between Confirmed TB (blue) and Unlikely TB (orange) groups (p-value < 0.05). In the heatmaps, Z-scores above 0 are shown in red and below 0 in blue.

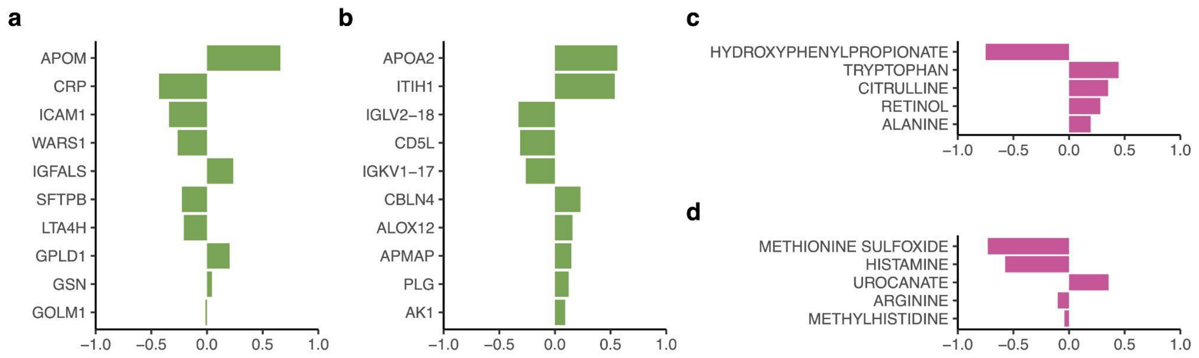


***Supplementary Figure 4.*** Feature contribution using the multi-omics integration approach DIABLO in the mixOmics package. (a, b) All contributing proteins on components 1 and 2 respectively, and (c, d) All contributing metabolites on components 1 and 2. In each plot, the x-axis indicates the loading factor of variables contributing to each component.

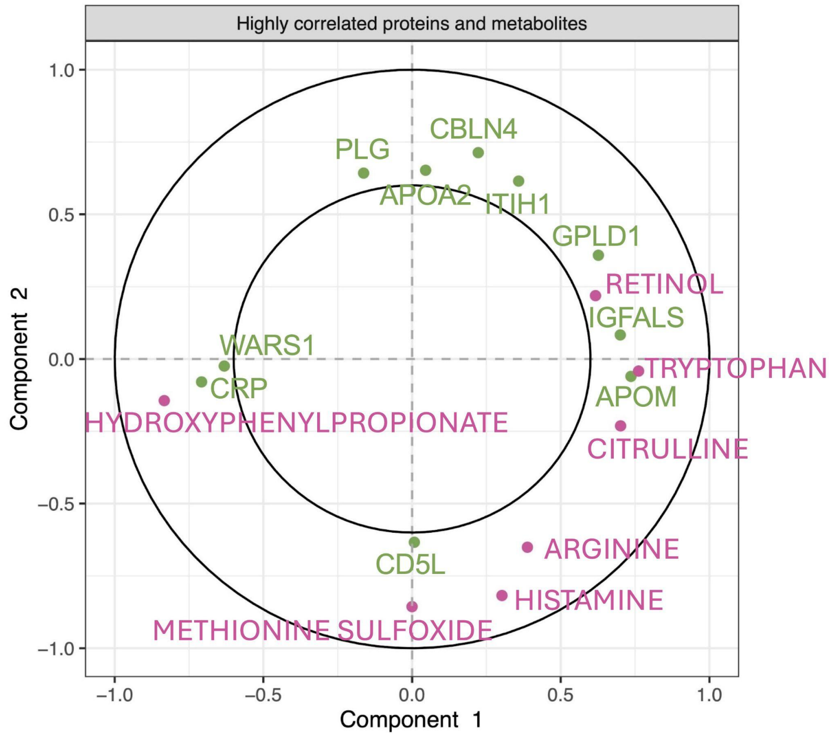


***Supplementary Figure 5.*** Variable correlation plot (varPlot) of proteins and metabolites associated with the two components identified using mixOmics. Each green point represents a protein, and each pink point represents a metabolite. The position of each point reflects its correlation with Component 1 (x-axis) and Component 2 (y-axis). The region of high correlation (0.6 to 1) is indicated by two black circles. This plot emphasizes the features that contribute most to the separation between Confirmed TB and Unlikely TB groups.

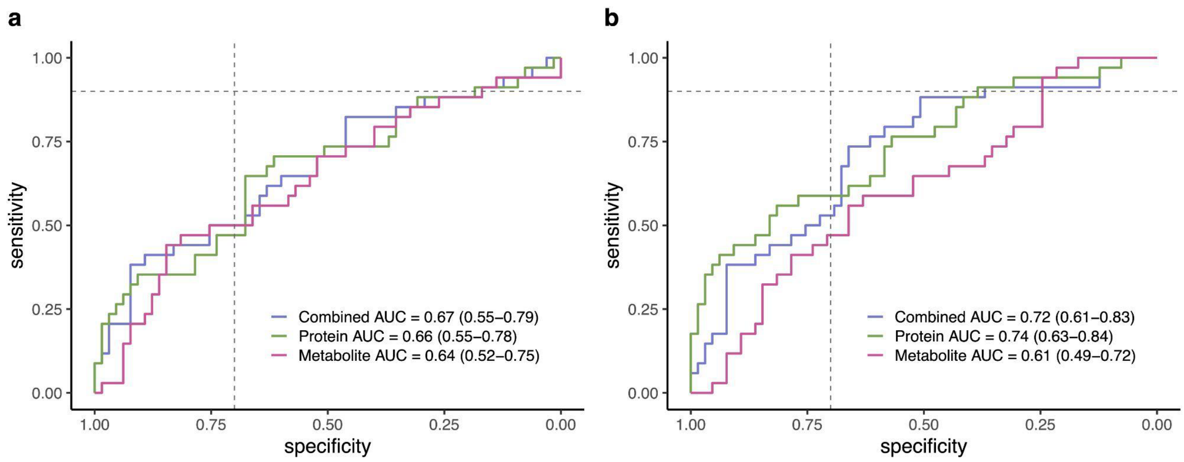


***Supplementary Figure 6*.** **(a)** ROC curves for models using all contributing proteins and metabolites to latent components from mixOmics (20 proteins and 10 metabolites). (b) ROC curves for models using highly correlated proteins and metabolites (10 proteins and 7 metabolites). AUC values are included for each model. The two dashed lines in two plots represent the target product profile (TPP) criterias for a TB triage test, defined by a sensitivity above 90% and a minimum specificity of 70%.

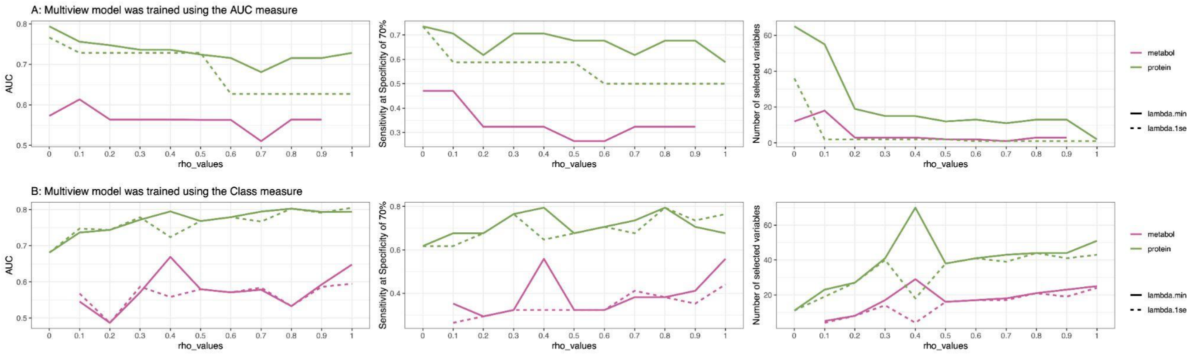


***Supplementary Figure 7.*** Performance comparison of the multiview model trained using the AUC measure (A) and class labels (B). Each panel includes three plots showing AUC, sensitivity at 70% specificity, and the number of selected features across different rho values (ranging from 0 to 1). The green and pink lines represent results for the proteomics and metabolomics datasets, respectively. Solid lines indicate performance using the less regularized model (lambda.min), while dashed lines show results for the more regularized model (lambda.1se).

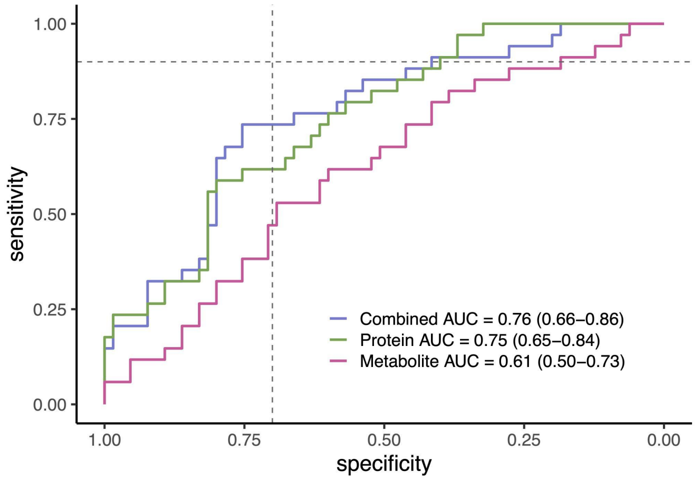


***Supplementary Figure 8.*** ROC curves for models using all selected proteins and metabolites (19 proteins and 18 metabolites) from multiview. AUC values with 95% confidence intervals are included for each model (proteins in green, metabolites in pink, and combined in purple).
